# Incorporating Genomic Sequences into Stochastic Transmission Modeling to Improve the Analysis of SARS-CoV-2 Transmission Dynamics

**DOI:** 10.64898/2025.12.11.25342070

**Authors:** Toni Gui, Ira Longini

## Abstract

The recent SARS-CoV-2 pandemic has highlighted the growing importance of infectious disease analysis. An accurate and robust model can empower public health leaders to make timely decisions on social distancing and vaccination policies, thereby reducing the number of cases, hospitalizations and deaths. However, the emergence of new variants and subvariants can significantly alter the transmissibility, immune escape capacity and virulence of the pathogen in a short time, making the number of cases, hospitalizations and deaths difficult to predict. To enhance the timeliness and accuracy of forecasting, SARS-CoV-2 sequencing data can be utilized. These data constitute a vast and continuously growing resource, with millions of sequences collected and reported over the past few years. By incorporating the evolution of SARS-CoV-2 virus into classic transmission models, we conclude that genomic data is crucial for capturing trends in epidemiological data when new variants and subvariants emerge, leading to the development of a more reliable model and enhancing our knowledge of transmission dynamics and control.

## 1 Introduction

The pandemic of SARS-CoV-2, since 2019, has drawn wide attention and posed unprecedented challenges to global public health. In addition, other infectious diseases, such as influenza and HIV, have been extensively studied for decades to prevent transmission and reduce mortality, yet research is still ongoing. To understand the underlying mechanism of these infectious diseases, it is essential to investigate the evolution and spread of the pathogens. New developments in data science for high-throughput sequencing are facilitating genetic data collection and optimizing data processing, potentially revolutionizing infectious disease epidemiology. However, how to efficiently utilize these high-dimensional sequencing data remains challenging, as they only offer raw genetic information and integrating them with other data types, such as epidemiological data, can be complex.

The field of phylodynamics brings evolutionary models and epidemiological processes together (Volz et al., 2013). Some earlier work jointly inferred transmission dynamics and phylogenies to study hepatitis C (Kühnert et al., 2014). Bayesian phylogenetic frameworks have been used to link genetic divergence to epidemic dynamics in structured populations (Kühnert et al., 2016). More recently, phylodynamics approaches allow for joint inference of epidemic parameters and evolutionary histories and have been applied to study early outbreaks of SARS-CoV-2 (Vaughan et al., 2024, Attwood et al., 2022). In addition, methods that combine particle filtering and phylogenetics attempt to jointly infer the incidence trend and the phylogenetic tree structure (Vaughan et al., 2019). Comparative studies of genomic epidemiological models show that integrating sequence data can provide more accurate inference than classical models along (Cárdenas et al., 2022). Yet challenges in phylodynamic inference remain, such as computational intensity, sampling biases, evolutionary model mis-specification, and the complexity of linking transmission events to phylogenetic divergence (Frost et al., 2015, Baele et al., 2016). Another study shows that integrating pathogen phylogenies derived from sequencing data with epidemiologic information within a survival-analysis framework can improve the precision of estimated transmission parameters, as shown through its application to the 2001 foot and mouth disease outbreak in the United Kingdom (Kenah et al., 2016).

Many existing integrative methods are computationally intensive, and require restrictive assumptions that can limit the applications in applied settings. Moreover, reconstructing full phylogenies can introduce additional complications, as some assumptions used to build the phylogenetic tree may not accurately reflect the real system. Simpler evolutionary indices that can be directly plugged into compartmental models may be preferred, as they allow genomic information to shape model parameters.

The Partially Observed Markov Process (POMP) framework provides a powerful foundation for modeling dynamic systems when key components are unobserved. It has been widely used in applied epidemiological modeling across various pathogens, including measles (Bretó et al., 2009), and cholera (A. King et al., 2008). The R package pomp was introduced in 2016, and it has been widely used in applied epidemiological modeling (A. A. King et al., 2016). For example, a study developed a forecasting model that combined hospital admissions and mobility data to monitor COVID-19 transmission and the healthcare demand in near real time (Fox et al., 2022).

In this work, we propose a novel index “sequencing distance” derived from genomic sequences that quantifies the rate of pathogen evolution. We describe how to incorporate this index directly into the compartmental modeling framework as a dynamic driver of key parameters: transmissibility, virulence, and immune escape. The model is implemented using the pomp package in R. Using both simulation experiments and real-world data, we show that incorporating the genomic index leads to notable improvements in model fit and predictive performance compared to models that do not apply genomic information. We also discuss limitations, computational considerations, and extensions such as age structure, vaccination, and behavioral data in more complex settings.

## 2 Materials and Methods

### 2.1 Data Resource and Processing

The genetic data used in our analysis is based on 10,000 complete SARS-CoV-2 sequences with high coverage collected in San Diego County between 2020-03-04 and 2022-02-01, along with epidemiological data for confirmed cases, hospitalizations and deaths between 2020-02-14 and 2022-02-01 posted on the official website of San Diego County. The sequences are aligned using ViralMSA, with SARS-CoV-2 WIV04 used as the reference sequence. The spike protein sequences are extracted from the aligned sequences and translated into amino acid sequences.

We define an index, the “weekly sequencing distance”, to measure the speed of evolution in the pathogen over time. The index value is obtained through the following steps. First, for each pair of spike protein sequences, we count the number of single amino acid polymorphisms (SAPs). Second, for each sequence *i*, we calculate the average of the highest 20 SAP counts from sequences collected within the previous 30 days, denoted as *d*_*i*_. Third, we average *d*_*i*_ for the sequences collected each week *t*, denoted as *a*_*t*_. Due to a lack of sequence collection at certain times, 15 weeks have missing averaging data. We assign values to those weeks using a spline, based on the nearest preceding and following weeks that have non-NA values; if no such week exists, we assign a value of 0. Finally, with the notation *SD*_*t*_ representing the sequencing distance for week *t*, we have

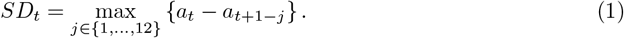

These sequence distances over time are shown in Figure 1.

**Figure 1:**
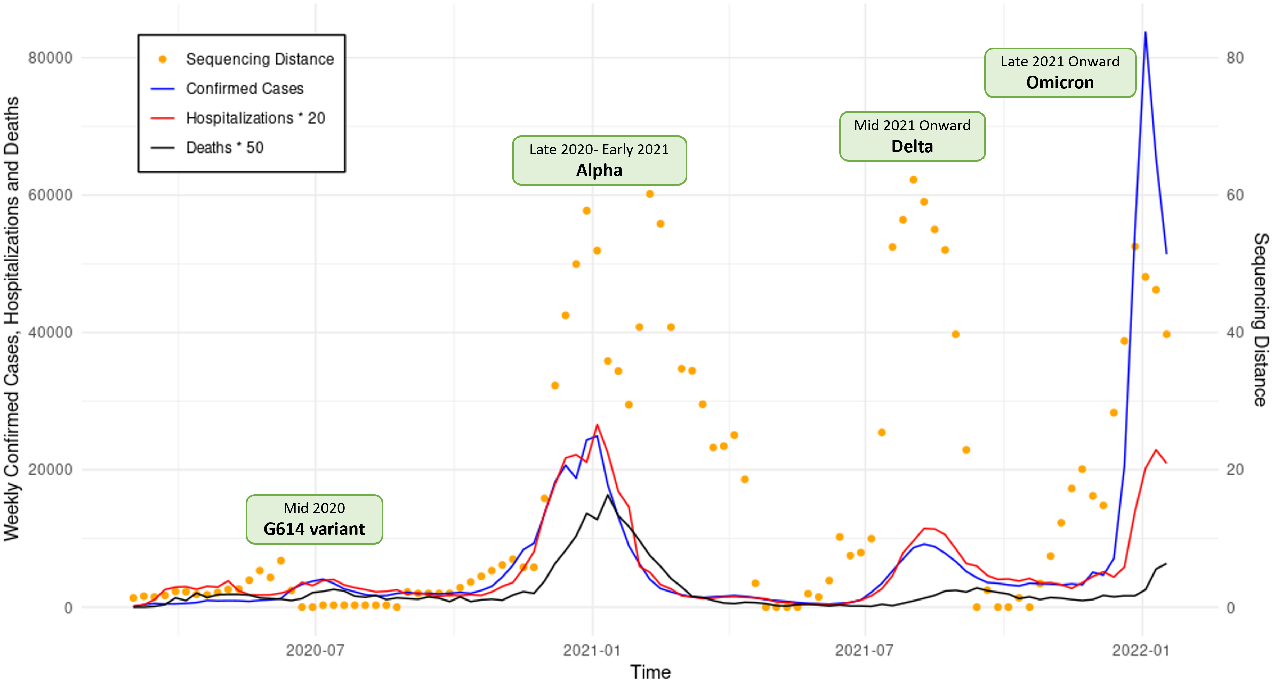
Weekly confirmed cases, hospitalizations, deaths and sequencing distance data in San Diego County. Also, the appearance of named virus variants.

When the sequencing distance becomes larger than more or less 10, the circulating virus tends to be labeled as a new variant: such as Alpha, Delta, and Omicron, as shown in Figure 1. We see, also, marked increases in the number of confirmed cases, hospitalizations, and deaths, once the sequencing distance becomes greater than 10.

### 2.2 Stochastic Transmission Model

We built an S-I-R-S mathematical model with the following compartments: susceptible (*S*), symptomatic infectious (*I*^*Y*^ ), asymptomatic infectious (*I*^*A*^), recovered (*R*), hospitalization (*H*), and deceased (*D*). This stochastic model is built on a discrete state space and in discrete time. The total size of the population at the beginning is *N* . We assume that in each period of time, a small random number of asymptomatic infected cases are imported from outside the population. This assumption ensures that the number of infected cases does not stay at zero if they reach zero. The model structure is shown in Figure 2.

**Figure 2:**
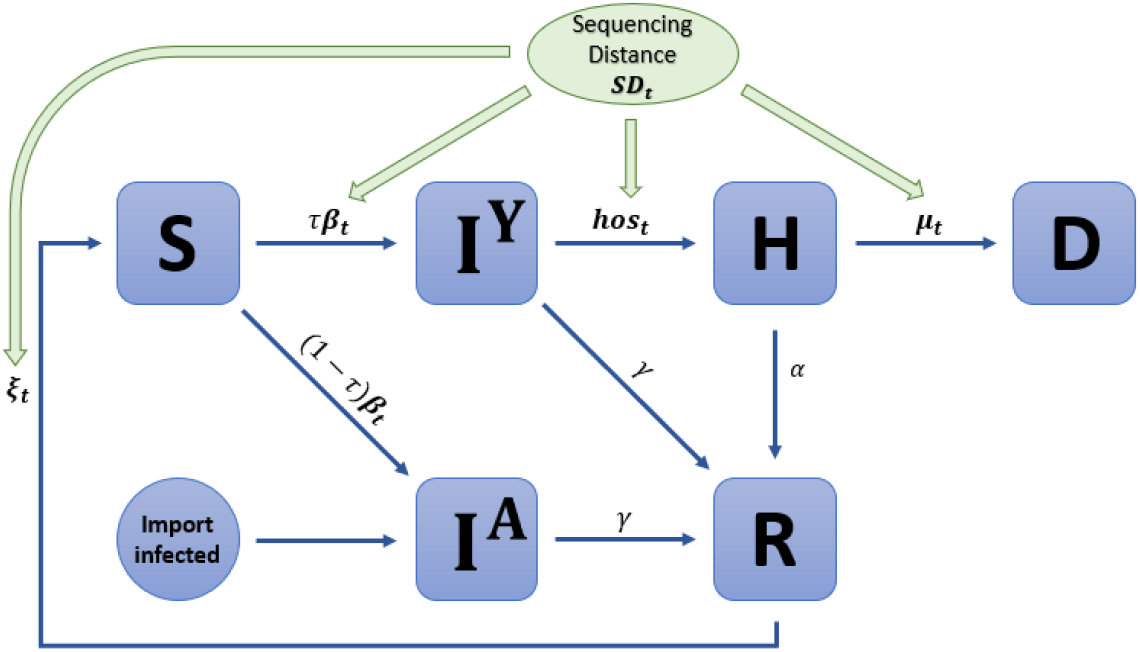
The compartmental model.

We let *β*_*t*_, *ξ*_*t*_, *hos*_*t*_ and *μ*_*t*_ be the time-dependent transmission rate, immunity loss rate, hospitalization rate and mortality rate, respectively. These rates vary over time as the dominant variant changes, with each variant exhibiting different values for each rate. We define the rates as functions of the average sequencing distance as follows:

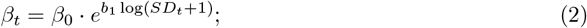

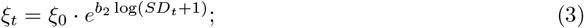

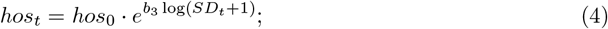

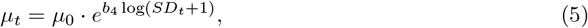

where *SD*_*t*_ is the sequencing distance defined in (1).

The transitions between compartments are governed by *τ* -leap method, with the parameters in Table 1.

**Table 1:**
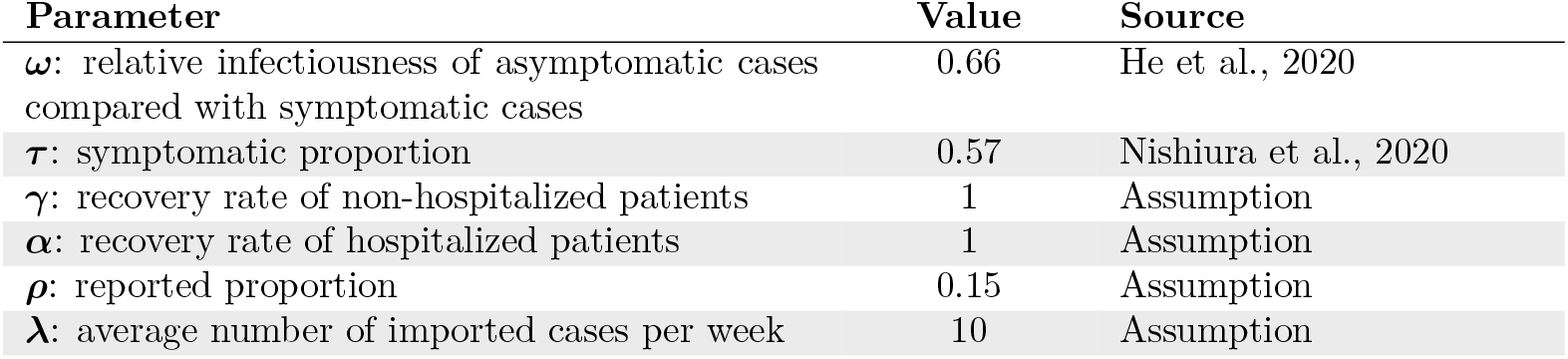
Parameters and Values.

| Parameter | Value | Source |
| --- | --- | --- |
| $\omega$: relative infectiousness of asymptomatic cases compared with symptomatic cases | 0.66 | He et al., 2020 |
| $\tau$: symptomatic proportion | 0.57 | Nishiura et al., 2020 |
| $\gamma$: recovery rate of non-hospitalized patients | 1 | Assumption |
| $\alpha$: recovery rate of hospitalized patients | 1 | Assumption |
| $\rho$: reported proportion | 0.15 | Assumption |
| $\lambda$: average number of imported cases per week | 10 | Assumption |

The deterministic dynamics of the system are governed by the following ordinary differential equations:

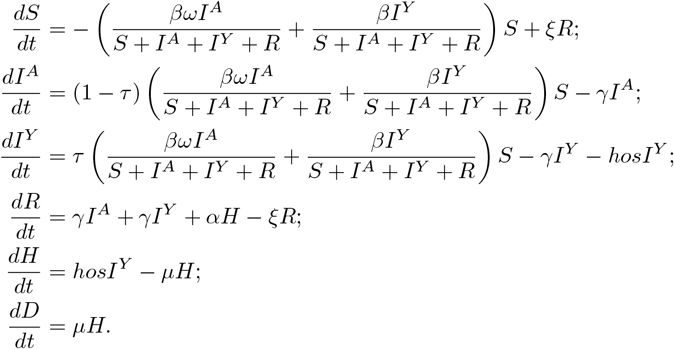

While the ordinary differential equations describe the deterministic dynamics of the system, such models fail to fully capture the randomness inherent in real-world transmission processes. Therefore, we employ a stochastic transmission model, governed by the following equations.

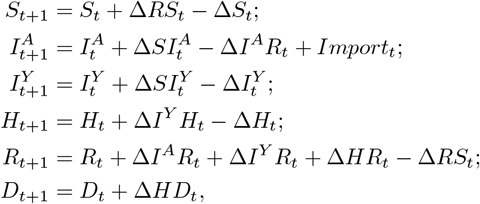

where

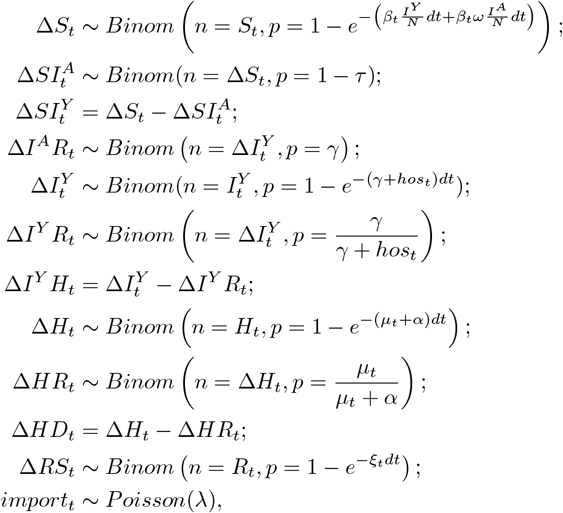

with the initial conditions

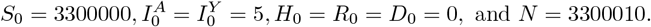

The notation *Binom*(*n, p*) and *P oisson*(*λ*) represent, respectively, a binomial distribution with *n* trials and success probability *p*, and a Poisson distribution with rate *λ*.

We assume that all reported data follow Poisson distributions. Let *RC*_*t*_, *RH*_*t*_ and *RD*_*t*_ be the number of confirmed cases, hospitalizations and deaths at time *t*, respectively. Due to a shortage of testing supplies, reporting resources and the presence of asymptomatic cases, only a proportion of the infections are reported. Suppose that

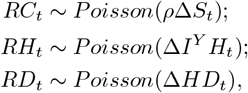

where *ρ* represents the proportion of infections reported.

Let ***θ*** denote the parameter vector

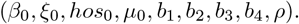

Let ***X***_***t***_ denote the state vector

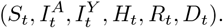

Let the lowercase form of each time-dependent random variable denote its realized value at the corresponding time. Then the likelihood function is

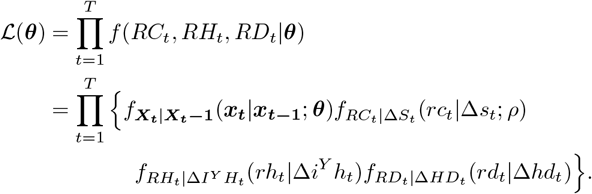

Given the assumed value of *ρ*, the log-likelihood can be computed using the particle filter for the estimation of the eight parameters.

## 3 Results

### 3.1 Simulations

To evaluate the performance of the proposed estimation framework, assuming the model holds, we constructed a simulation study implemented via the pomp package. Parameter values were uniformly sampled from plausible epidemiological ranges shown below:

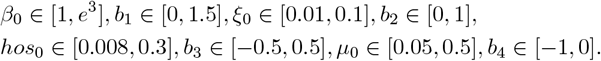

To evaluate the robustness of the method, 50 independent parameter sets were generated, and the model was assessed for its ability to recover or adapt to this variation accurately. Table 1 shows the mean error and 95% confidence interval coverage probability in the simulations. In all cases, the point estimates are nearly unbiased, which is indicated by the mean error squared being substantially smaller than the mean squared error itself. The 95% confidence interval coverage probabilities were close to 0.95, with only slight deviations. Thus, we are confident that the estimation procedure works well when the model is correct.

Figure 3 shows a simulated dataset of confirmed cases, hospitalizations and deaths, generated using a parameter set designed to produce trends that resemble those observed in real-world data.

**Figure 3:**
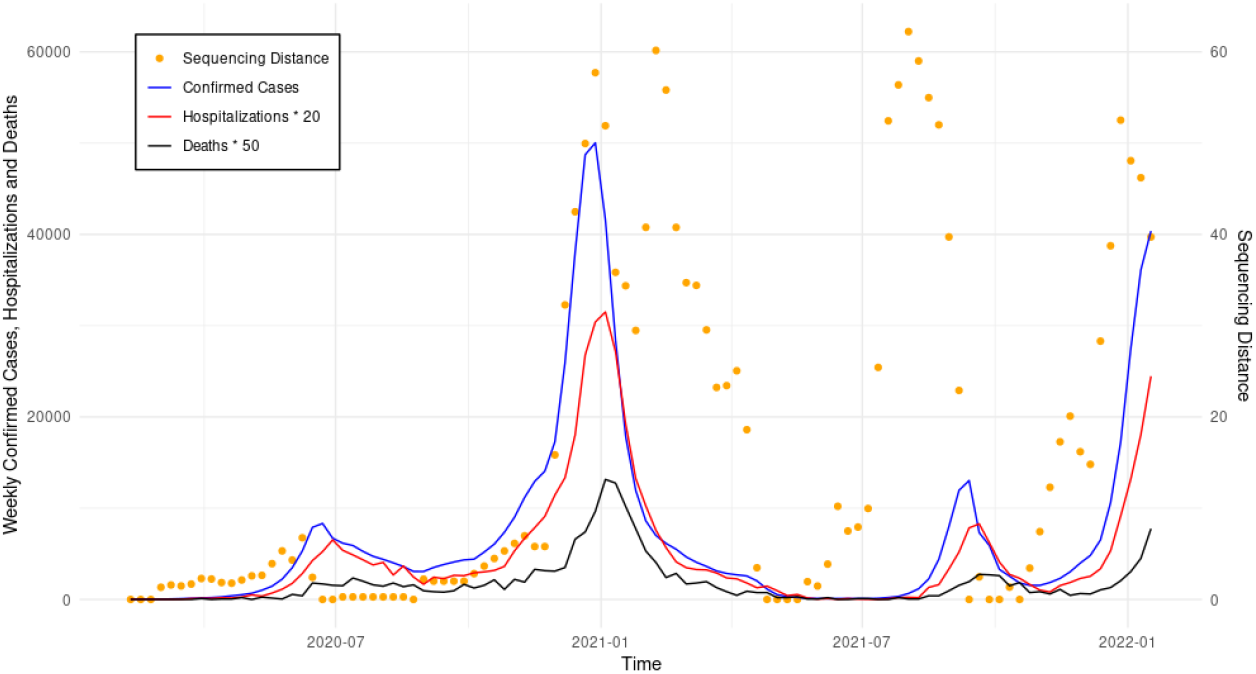
Simulated epidemic trajectories (confirmed cases, hospitalizations and deaths) and sequencing distance data over time.

Parameter estimates and their associated 95% confidence intervals were obtained using the pomp package. Table 3 summarizes the model parameters to be estimated, along with their maximum likelihood estimators and corresponding 95% confidence intervals.

**Table 2:**
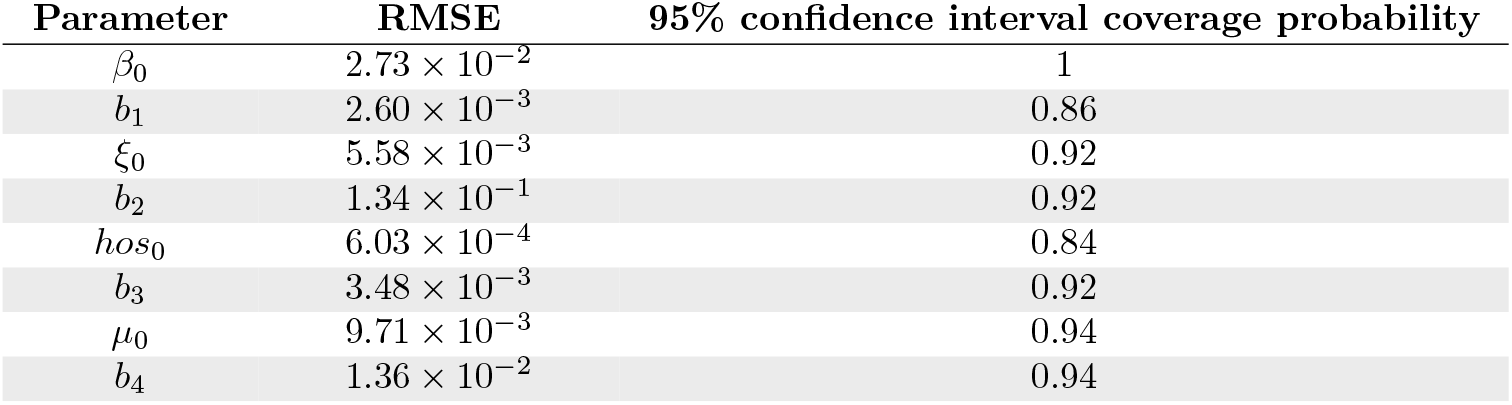
Statistical performance for the estimators.

**Table 3:**
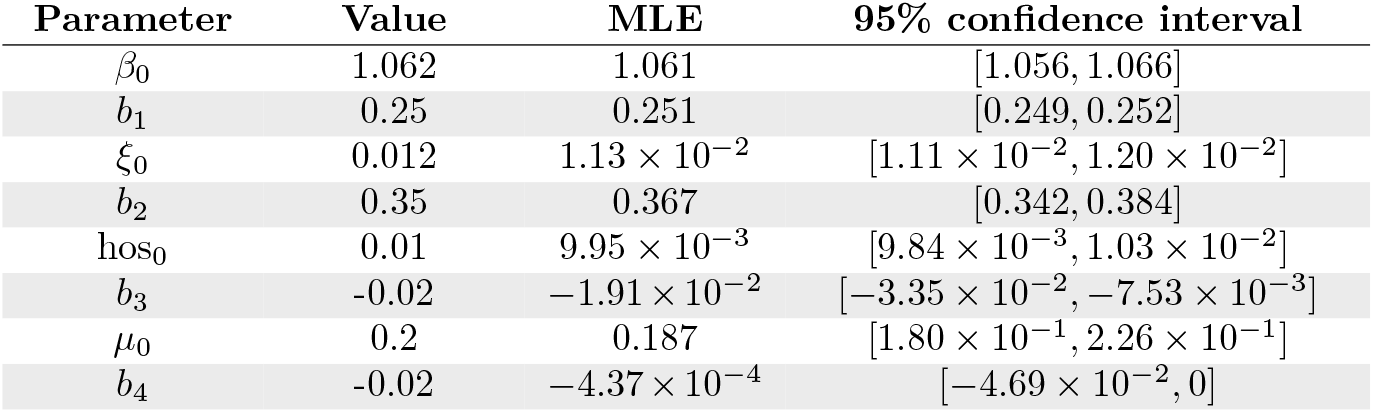
Summary of estimated parameters.

To visualize uncertainty, credible bands were generated from 2000 simulated trajectories by extracting the 2.5th and 97.5th percentiles at each time point. We see from Figures 4-6 that the model fits the simulated data well for cases, hospitalizations, and deaths.

**Figure 4:**
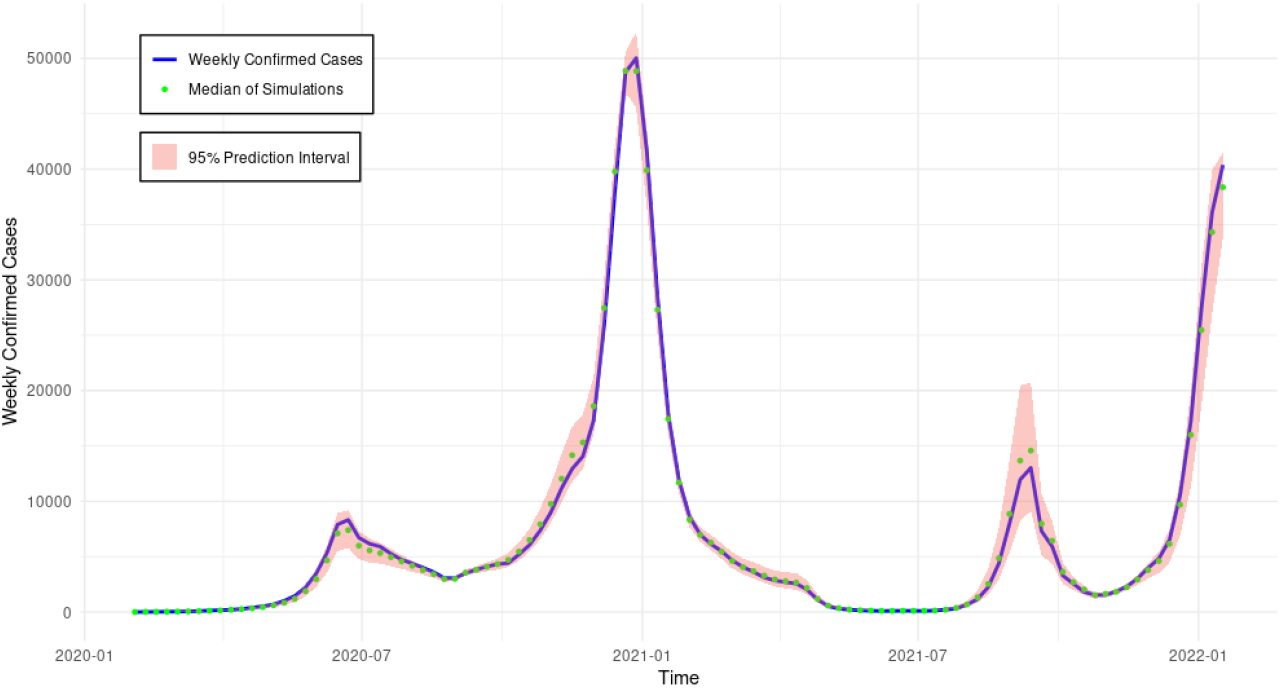
Weekly confirmed cases with median simulated values and its 95% credible band.

**Figure 5:**
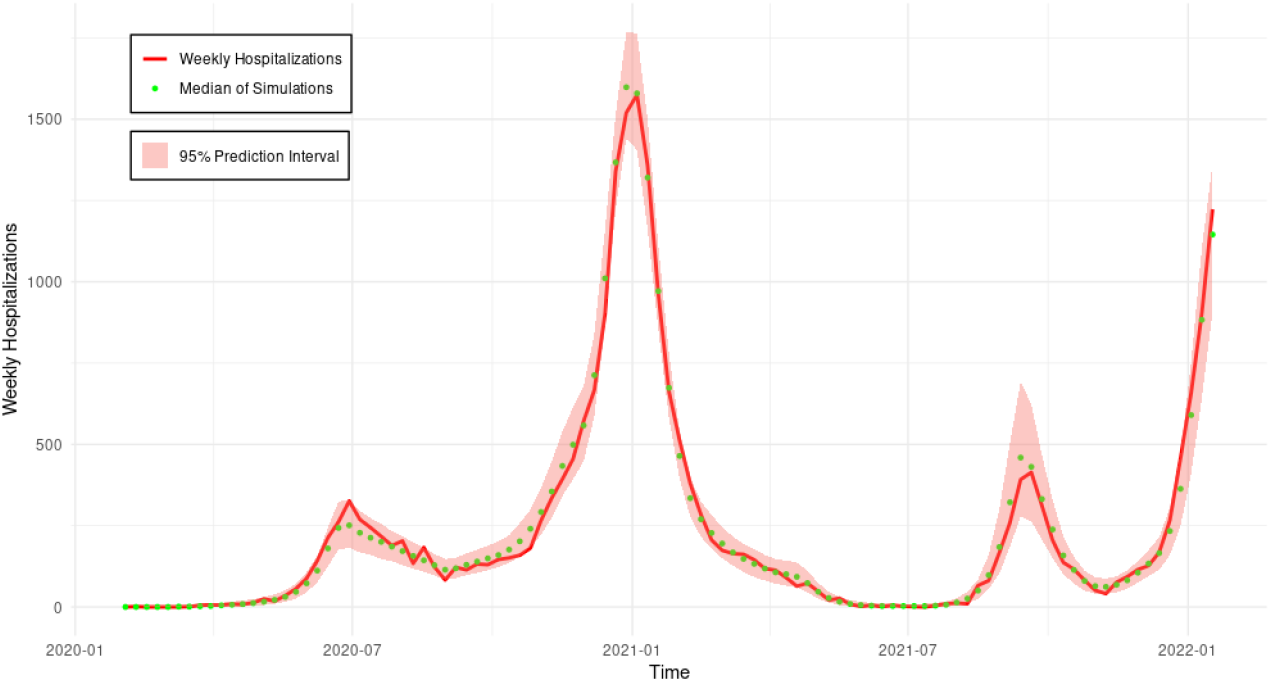
Weekly hospitalizations with median simulated values and its 95% credible band.

**Figure 6:**
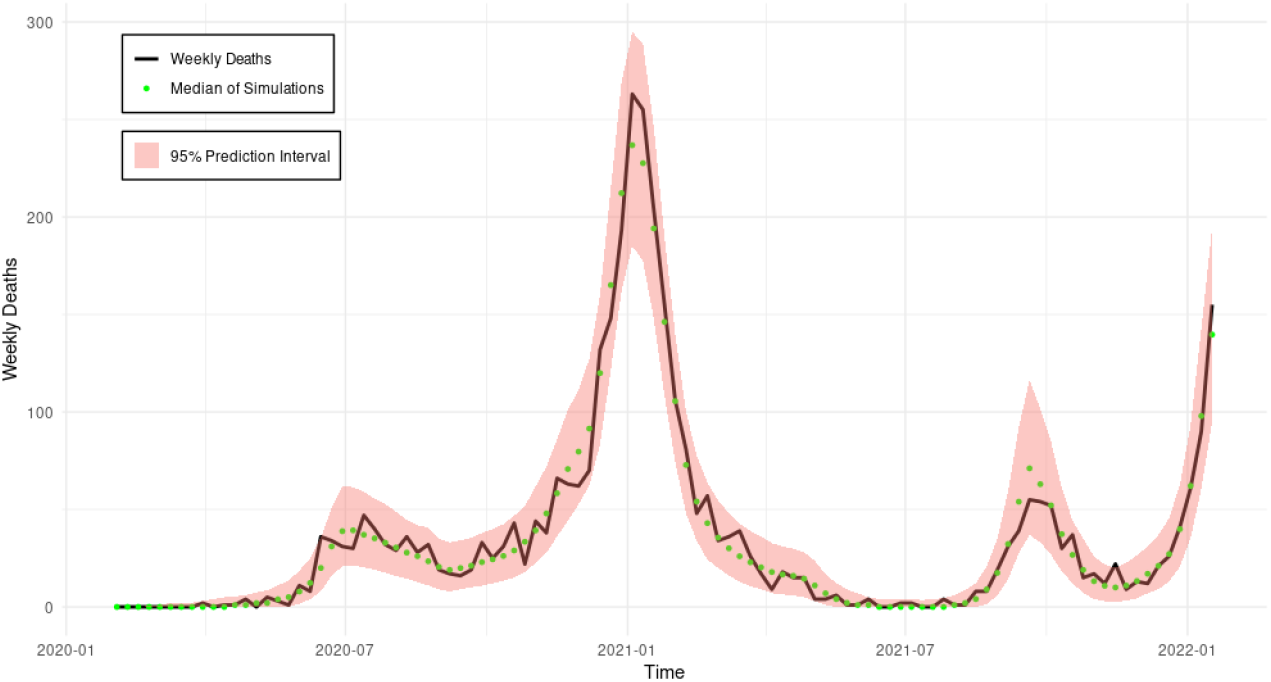
Weekly deaths with median simulated values and its 95% credible band.

### 3.2 Application to Real Data

Building upon the simulation study, which has demonstrated the model’s ability to recover parameters and capture expected epidemic dynamics under the assumption that the model structure correctly reflects the underlying system, we now apply the approach to real-world data to evaluate its empirical performance and applicability.

The fitting results are shown in Figures 7, 8, and 9, illustrating the alignment between the observed data and the model predictions. The uncertainty band are noticeably wider than those in the simulation results, mostly due to greater uncertainty in parameter estimates when fitting to real-world data. From Figures 7-9, we see that the model was able to pick up the timing of the increases well, but not the magnitude of the increases. This was due to the fact that we are using a simple model that does not include the effects of lockdowns, vaccination, and changes in reporting.

**Figure 7:**
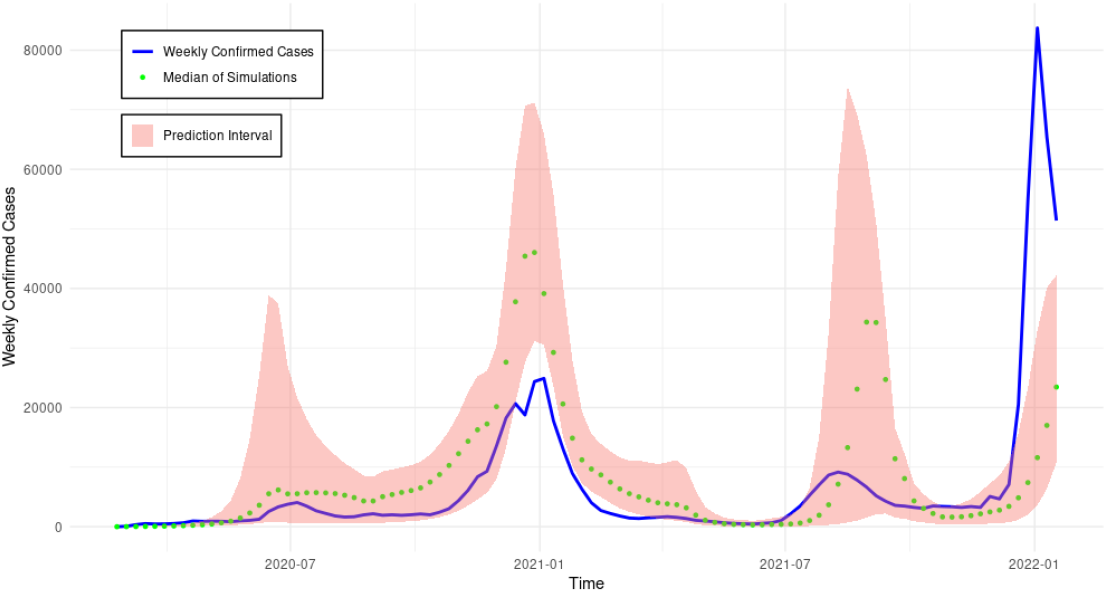
Weekly confirmed cases in San Diego County with median simulated values and uncertainty band.

**Figure 8:**
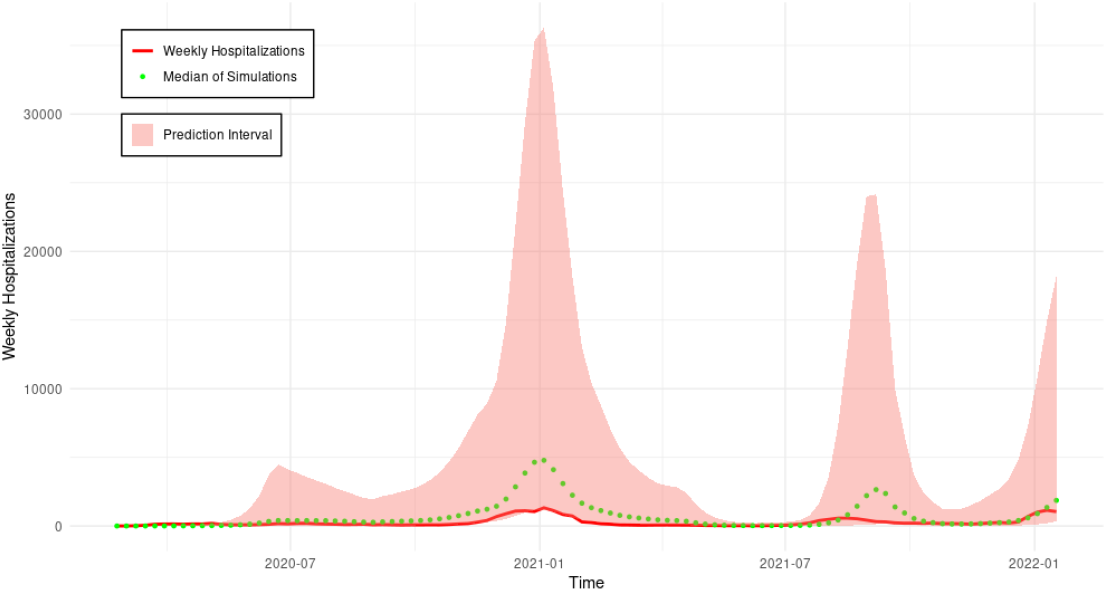
Weekly hospitalizations in San Diego County with median simulated values and uncertainty band.

**Figure 9:**
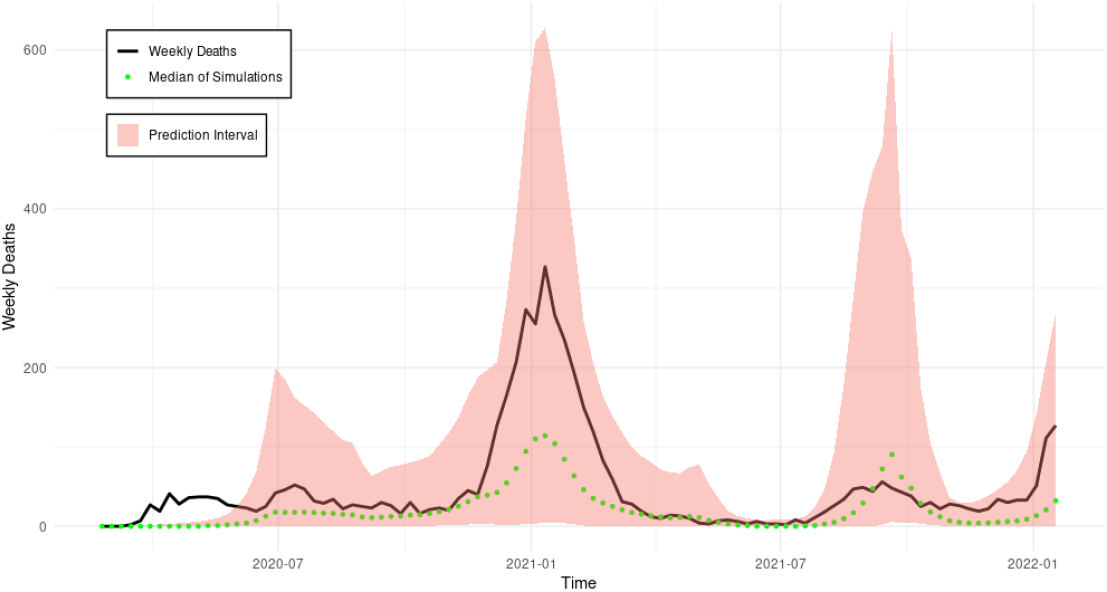
Weekly deaths in San Diego County with median simulated values and uncertainty band.

## 4 Discussion

In this study, we investigated how incorporating genomic data into stochastic compartmental models can enhance the model’s fit to epidemiological data when the pathogen undergoes evolution. The sequencing distance we defined offers an accessible approach to quantifying the rate of pathogen evolution. Instead of relying on raw genomic data, this index can be directly integrated into the parameter functions of compartmental models, enabling incorporation of the evolutionary rate into transmission modeling.

Our simulation results indicate that, given the current model setup, the integration of genomic data significantly improves the performance of model fitting. This conclusion also holds in real-data analysis. It is well known that the dynamics generated by the traditional S-I-R-S compartmental model typically exhibits an initial high incidence peak followed by smaller and smaller waves, *i*.*e*., damped oscillations towards an asymptotic fixed point. However, such behavior does not capture the complexity and variable patterns observed in real-world epidemiological data. Our real-data fitting results show that incorporating genomic data substantially improves the model’s ability to fit real-world observations. The model successfully captures key epidemiological trends, particularly during the emergence of notable variants or subvariants.

Although the mechanisms by which nucleotide sequences influence viral virulence and immune escape are undoubtedly complex and rooted in detailed biological processes, our method provides a simplified, reusable framework that does not rely on prior biological knowledge. This makes it more accessible for integration into traditional modeling pipelines, especially in settings where genomic data is available but domain expertise may be limited. Although the current trend of implementing machine learning methods in infectious disease modeling offers strong predictive capabilities, they often lack interpretability, limiting their usefulness for understanding transmission dynamics. In contrast, our model is better suited to generate meaningful epidemiological insights.

The simulation results under the current model appear reasonably consistent and show that with the consideration of genomic data, the data fitting does work better than the model without it, which indicates that the model can capture the epidemiological dynamics under simplified assumptions. However, achieving model convergence when applied to real-world data can be both challenging and time-consuming. Moreover, the model demonstrates suboptimal accuracy, suggesting that certain key components may not yet have been adequately captured. Future work will focus on extending the framework by integrating additional elements and mechanisms to improve empirical performance and more accurately reflect the underlying transmission dynamics. Specifically, the virulence and immune escape ability of SARS-CoV-2 have been shown to be more closely associated with the ORF8 protein than with the spike protein. Therefore, using the sequencing distance derived from other proteins instead of solely the spike protein is likely to improve the model performance. Additionally, behavioral data has not been incorporated into the model, despite its significant influence on transmission dynamics. In particular, public health interventions such as mask mandates, social distancing and other policy measures will be considered in future extensions to capture human behavioral responses. Furthermore, next-phase modeling efforts should incorporate age structure and vaccination dynamics to better represent the complexity of real-world epidemic processes and enhance the model’s applicability to public health interventions. We also plan to incorporate mechanisms such as time-varying transmission rates and heterogeneity in contact structures into future modeling efforts. These additions will aim to capture more realistic population-level behaviors and improve the model’s ability to reproduce complex patterns observed in real-world data.

## Data Availability

All data used are publicly available.

## 5 Acknowledgement

This research was partially funded by the CDC grant NU38FT000013, entitled EPISTORM: Center for Advanced Epidemic Analytics and Predictive Modeling Technology, and by the WHO R&D Blueprint for Epidemics.

## Notes

### Competing Interest Statement

The authors have declared no competing interest.

